# The impact of the COVID-19 pandemic on health service utilisation in Sierra Leone

**DOI:** 10.1101/2021.04.12.21255327

**Authors:** Stephen Sevalie, Daniel Youkee, Alex J van Duinen, Emma Bailey, Thaimu Bangura, Sowmya Mangipudi, Esther Mansaray, Maria-Lisa Odland, Divya Parmar, Sorie Samura, Diede van Delft, Haja Wurie, Justine I Davies, Håkon Bolkan, Andrew JM Leather

## Abstract

**Introduction:** The COVID-19 pandemic has adversely affected health systems in many countries, but little is known about effects on health systems in sub-Saharan Africa. This study examines the effects of COVID-19 on health service utilisation in a sub-Saharan country, Sierra Leone.

**Methods:** Mixed-methods study using longitudinal nationwide hospital data (admissions, operations, deliveries and referrals), and qualitative interviews with healthcare workers and patients. Hospital data were compared across Quarters (Q) in 2020, with day 1 of Q2 representing the start of the pandemic in Sierra Leone. Admissions are reported in total and disaggregated by sex, service (surgical, medical, maternity, paediatric), and hospital type (government or private not for profit). Referrals in 2020 were compared with 2019, to assess whether any changes were the result of seasonality. Comparisons were performed using student’s t test. Qualitative data were analysed using thematic analysis.

**Results:** From Q1-Q2, weekly mean hospital admissions decreased by 14.7% (p=0.005). Larger decreases were seen in male 18.8%, than female 12.5% admissions. The largest decreases were in surgical admissions, a 49.8 % decrease (p<0.001) and medical admissions, a 28.7% decrease (p=0.002). Paediatric and maternity admissions did not significantly change. Total operations decreased by 13.9% (p<0.001), whilst caesarean sections and facility-based deliveries showed significant increases, 12.7 % (p=0.014) and 7.5% (p=0.03) respectively. In Q3 total admissions remained 13.2% lower (p<0.001) than Q1. Mean weekly referrals were lower in Q2 and Q3 of 2020 compared to 2019, suggesting findings were unlikely to be seasonal. Qualitative analysis identified both supply-side factors, prioritisation of essential services, introduction of COVID-19 services and pausing elective care, and demand-side factors, fear of nosocomial infection and financial hardship.

**Conclusion:** The study demonstrated a decrease in health service utilisation during Covid-19, the decrease is less than in other countries during COVID-19 and less than reported during the Ebola epidemic.

**What is already known?:**

- During the Ebola epidemic, Sierra Leone experienced drastic reductions in health service utilisation, that are thought to have led to high mortality.
- Reductions in healthcare utilisation have been reported in other countries due to the COVID-19 pandemic, however little is known about the effects of the pandemic on healthcare utilisation in sub Saharan Africa, including Sierra Leone.

**What are the new findings?:**

- Healthcare utilisation in Sierra Leone decreased modestly during the first wave of the COVID-19 pandemic.
- Decreases in hospital admissions were less than those seen during Ebola and less than decreases seen globally.
- The largest reductions were seen in adult medical and surgical services, populations covered under the free healthcare act including maternal and child (under 5 years) health were more resilient.

**What do the new findings imply?:**

- The minimal reduction in service utilisation suggest that lessons have been learnt in protecting essential health services during outbreaks.
- Similar patterns of decreases in healthcare utilisation from COVID-19 to Ebola, should inform future preparedness and outbreak response planning.
- The resilience of services covered by the free healthcare initiative supports the argument for Universal Health Coverage in Sierra Leone.

## Introduction

The COVID-19 pandemic is the largest outbreak of an infectious disease in recent history. Sierra Leone reacted quickly to the threat, implementing policies to contain the pandemic. As of 30/03/2021, Sierra Leone had 3970 confirmed cases of COVID-19 and 79 recorded deaths(1). Evidence from past epidemics(2) and initial reports from the current pandemic, suggest that the greater threat in sub-Saharan Africa (sSA) countries, including Sierra Leone, may well be the indirect effects of COVID-19 on access and delivery of essential health services(3).

Thus far, there has been limited research on the impact of COVID-19 on essential health services in sSA, including Sierra Leone. A systematic review published in 2021, of the impact of COVID-19 on healthcare utilisation worldwide, found no eligible studies from sSA and only four studies from non-African Low and Middle Income countries (LMICs)(4). In rural South Africa, a single centre interrupted time series analysis, found no significant change in total admissions during 2020, but did find significant changes between subgroups of admissions(5). Hospital level data from South Africa and Nigeria, documented that antenatal visits decreased whereas evidence was mixed for facility-based deliveries and caesarean sections(6). Interviews of community stakeholders from Kenya and Nigeria, found that stakeholders perceived a reduction in access to healthcare during COVID-19 lockdowns; perceived barriers were cost, reduced availability of transport and fear of infection(7). Other articles concerning sSA present either modelled data(8), opinion or recommendations(9).

Outside of sSA, evidence has shown the impact of the pandemic on all aspects of care(4). For example, Pakistan reported a 52.5% decline in the daily average total number of vaccinations administered during their lockdown compared to baseline(10). A prospective observational study from Nepal reported facility-based deliveries reduced by 51.4 %, with a corresponding increase in maternal and infant mortality(11). A nationwide study in China found that total healthcare expenditure and utilisation declined by 37.8% and 40.8%, respectively during the worst phase of the outbreak(12). The impact of the pandemic on surgical services is indicated in an expert elicitation exercise involving 190 countries. It was estimated that 2 367 050 operations were cancelled per week during the 12 weeks of maximum COVID-related disruption in 2020(13). Others have found similar reductions in surgical activity(14).

Sierra Leone has recent experience of the impact of a viral pandemic on essential health services. The 2013-2016 Ebola epidemic caused a significant decline in both supply of, and demand for, essential health services in the region(2, 15). In Sierra Leone, a nationwide study demonstrated a 50% median reduction in inpatient admissions and a 41% median reduction in major operations performed during the Ebola epidemic compared to before(16). However, some services showed resilience, for example, caesarean deliveries increased in government hospitals, likely absorbing the effect of private hospital closures. A systematic review found that the largest decreases were seen for inpatient care and deliveries(15). The impact of Ebola on healthcare utilisation in Sierra Leone has been well studied, and provides a useful comparator to the impact of COVID-19. Through comparison of healthcare utilisation between the two outbreaks we can gain insights into health system resilience(17) during epidemics.

In this study, we aimed to determine the effects of the COVID-19 pandemic on health service utilisation by observing hospital admissions, surgical activity, and referral data from secondary and tertiary hospitals in Sierra Leone. We used qualitative method to understand the changes in demand for healthcare that contributed to healthcare utilisation patterns. We then describe the changes in the supply of health services, outlining the adaptive, planned health service reconfigurations and the unplanned service disruptions.

## Methodology

### COVID-19 Context

The first COVID-19 case in Sierra Leone was recorded on the 30^th^ March 2020 (Figure 1). Cases increased to a peak in June 2020 before steadily declining. Public health measures introduced in Sierra Leone were less strict than other Sub Saharan African countries(18). There were two 3-day national lockdowns between 5^th^ to 7^th^ April and 3^rd^ to 5^th^ of May 2020. Advice regarding social distancing and handwashing was widely disseminated, and mass gatherings, including religious ceremonies, were banned until further notice. Schools and colleges were closed, but shops and businesses remained open. A mandatory mask-wearing policy was introduced on 6^th^ July 2020. From 14^th^ April until 4^th^ July there was a ban on inter-district travel. From 22^nd^ March until 22^nd^ July 2020 international commercial air travel was suspended. These measures were reviewed and revised as the outbreak evolved in Sierra Leone.

**Figure 1:**
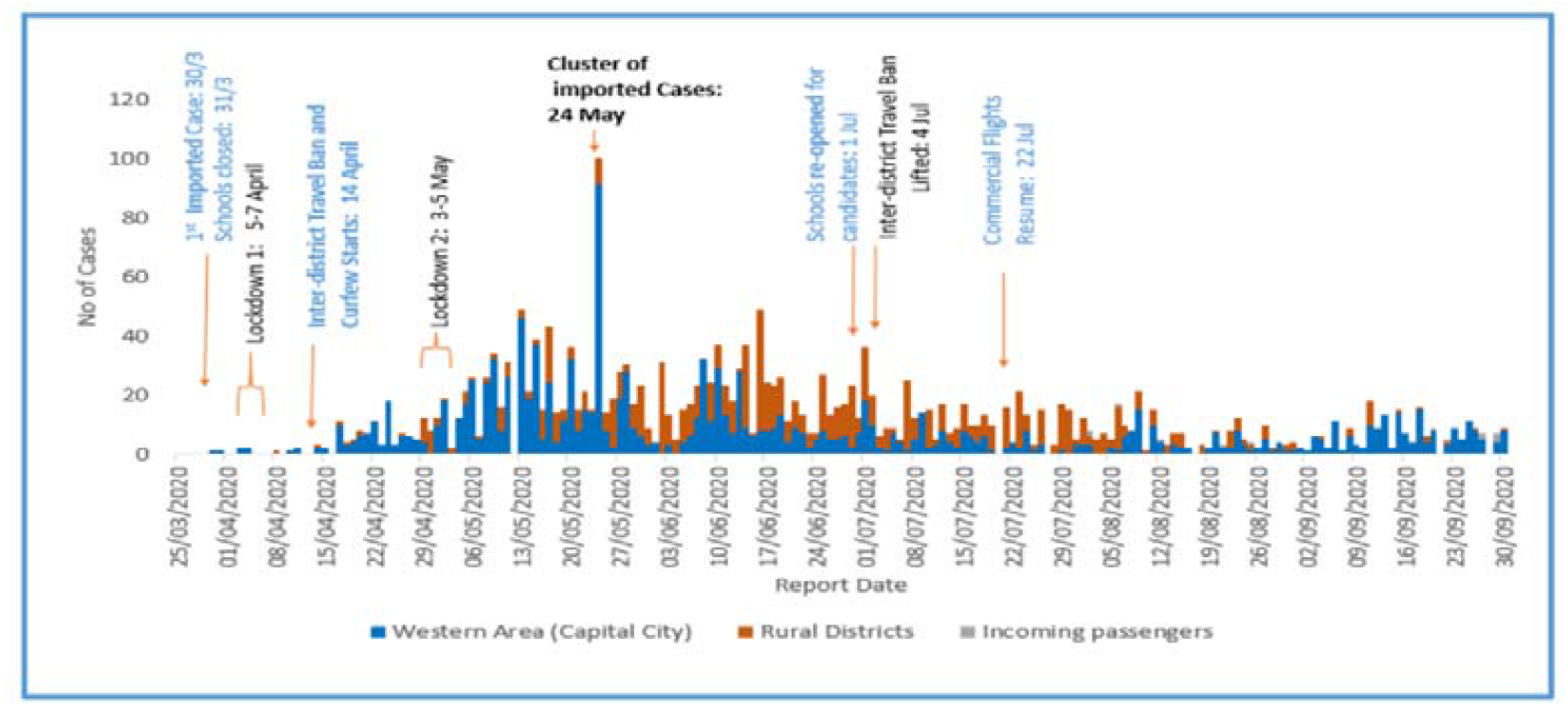
Daily COVID-19 cases in Sierra Leone as of 30th september 2020; Western Area includes Western Urban and Western Rural Districts; Rural Districts includes all other 14 Districts

We performed a mixed-methods study consisting of a retrospective study of nationwide hospital admissions, operations, and deliveries; a retrospective and prospective study of nationwide referrals; and a qualitative study using interviews with healthcare workers and patients from two districts.

Hospital data was collected from January 1^s^t 2020 until September 30^th^ 2020 by trained surgical assistant community health officers. The list of hospitals providing surgical services was sourced from a previous mapping study(19). Facilities which did not consent to data collection and facilities with known very low surgical volumes were excluded. Data were collected from the hospital admission book, the operation theatre book, and the maternity ward book. Variables collected reflected the number of inpatient admissions, hospital deliveries, and total number of operations performed in the operating theatre. Data were also collected on the number of caesarean sections and elective hernia operations as a proxy for emergency or elective surgical care provision respectively.

Nationwide referral data was accessed from the National Referral Service database at secondary and tertiary hospitals, from January 1^st^ 2019 until 30^th^ September 2020. These data on patients received at destination facilities from another facility are recorded by referral coordinators (RC) based at each hospital onto a standardised paper case report form and then transcribed into an EpiInfo™ datasheet. Data were extracted from this datasheet on the number of daily referrals to secondary or tertiary facilities. Data on admissions were not available for 2019, therefore the availability of referral data across two years allowed us to assess if any changes in admission data were due to seasonal variation. Neither hospital admission data nor referral data include information on COVID-19 admissions.

The primary outcome was the total number of admissions. Secondary outcomes included the number of hospital admissions disaggregated by sex, service (surgical, medical, maternity, paediatric), and hospital type (government or private not for profit). Other secondary outcomes were the total number of facility-based deliveries, operations, caesarean sections and hernia operations. Additional outcomes included the total number of referrals and the number of facility-based deliveries.

Referrals and admissions to facilities per week are described as a count shown graphically. The first COVID-19 case occurred on 30th March 2020(2), enabling a comparison of mean numbers of weekly admissions, operations, deliveries and referrals occurring in the first quarter (Q1) with those in the second (Q2) or third quarter (Q3) of the year. Differences between quarters are shown as a percentage. The average number of referrals per week in each quarter was compared between 2019 and 2020 using referral coordinator data. Comparisons were done using student’s T-test and 95% Confidence Intervals (Cis) calculated. Statistical analysis was performed in STATA v16, StataCorp™, SPSS™ and Microsoft Excel™.

The qualitative study used semi-structured interviews with health staff and patients. Five health facilities were purposely selected from two districts, Western Area Urban and Bo, to ensure a mix of rural and urban facilities covering different levels – tertiary and secondary hospitals and primary health units. In each facility, we interviewed two to three health staff and one patient. We interviewed staff based on their availability and as far as possible, staff from the surgical or maternity departments and those involved in the management of the facility. Patients were approached in the health facilities and if they agreed to be interviewed, were then interviewed later at their home or a convenient location. All respondents provided signed consent. Interviews were audio-recorded and conducted in English or Krio; Krio interviews were translated to English during transcription by trained bilingual Sierra Leonean qualitative researchers. Qualitative analysis was performed using Nvivo 12™ to identify common themes(20).

Ethical approval was granted by the Sierra Leone Ethical and Scientific Review Committee and the Regional Committee for Medical and Health Research Ethics in central Norway (2020/155388). Patients and the public were not involved in the design, or conduct, of our research.

## Results

A total of 60 hospitals were identified that performed surgery in Sierra Leone in 2017 (Figure 2), 20 of those were small private for-profit facilities with very low surgical volume and were not considered for inclusion in the current study(19). Out of the 40 eligible hospitals, 32 agreed to share data for this study, their geographic location and sector is described in table 1. In 2017, these 32 included hospitals performed 87.1% of the nationwide surgical volume(19). For the qualitative analysis we interviewed 12 staff and five patients.

**Table 1:**
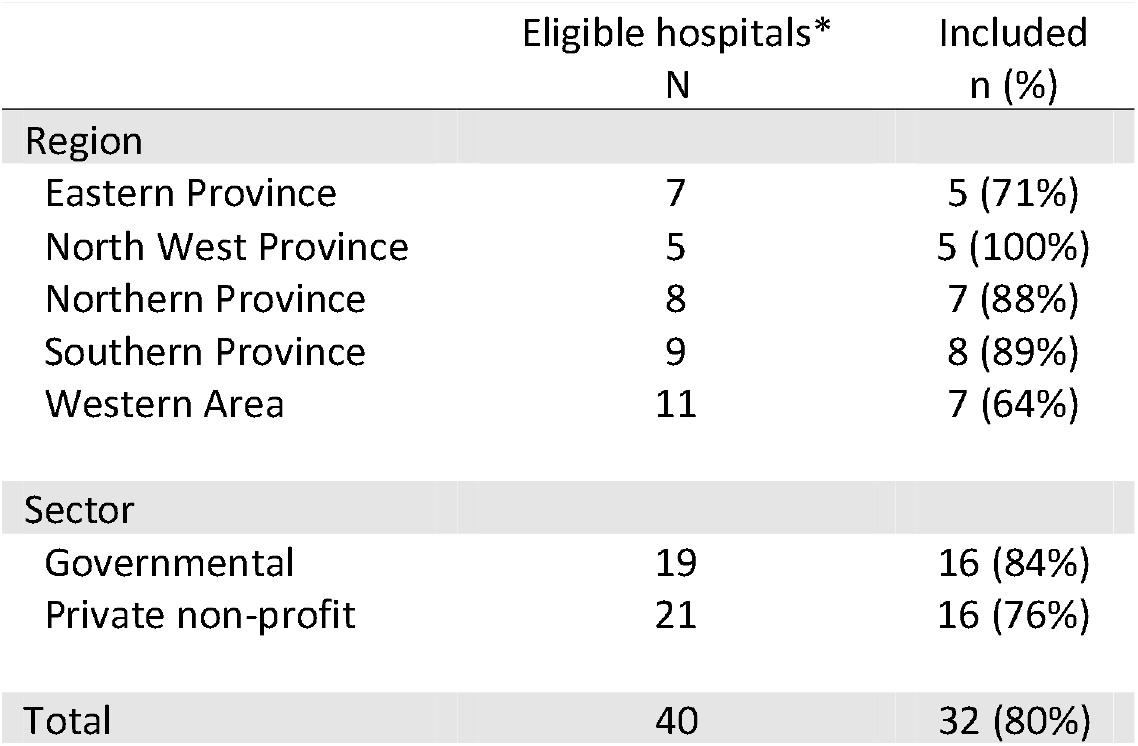
Hospital characteristics. *Governmental and private non-profit hospitals identified as providing surgery during 2017 mapping surgical activity in Sierra Leone(19).

|  | Eligible hospitals* | Included |
| --- | --- | --- |
|  | N | n (%) |
| **Region** |  |  |
| Eastern Province | 7 | 5 (71%) |
| North West Province | 5 | 5 (100%) |
| Northern Province | 8 | 7 (88%) |
| Southern Province | 9 | 8 (89%) |
| Western Area | 11 | 7 (64%) |
| **Sector** |  |  |
| Governmental | 19 | 16 (84%) |
| Private non-profit | 21 | 16 (76%) |
| **Total** | **40** | **32 (80%)** |

**Figure 2:**
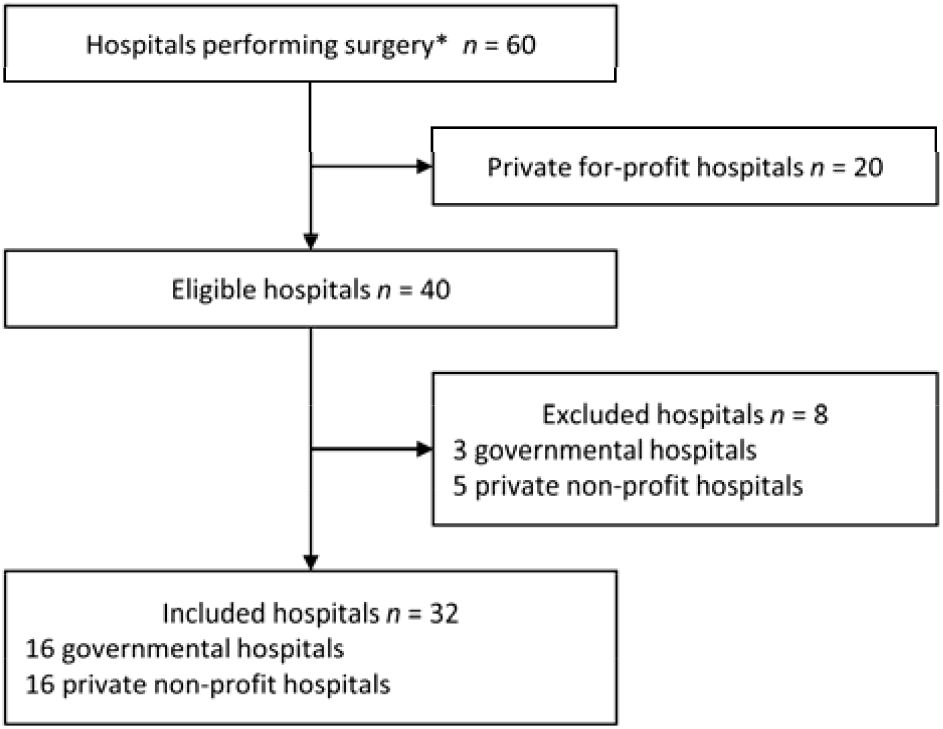
Hospital inclusion flowchart. n = number; *hospitals identified as providing surgery during 2017 mapping surgical activity in Sierra Leone(19).

Mean weekly hospital admissions in each quarter are shown in Table 2. From Q1 to Q2, there was a statistically significant decrease in nationwide mean weekly admissions from 2160 to 1842, a 14.7% decrease (p=0.005) (Figure 3a). Male admissions reduced from 753 to 611, a 18.8% decrease (p=0.004), and female admissions reduced from 1407 to 1231, a 12.5% decrease (p=0.009). Total mean weekly admissions remained low in Q3 compared to Q1, from 2160 to 1876, a 13.2% decrease (p<0.001). We found no significant recovery in Q3 compared to Q2, with total admissions Q3 to Q2, showing a 1.8% increase (p=0.715).

**Figure 3a:**
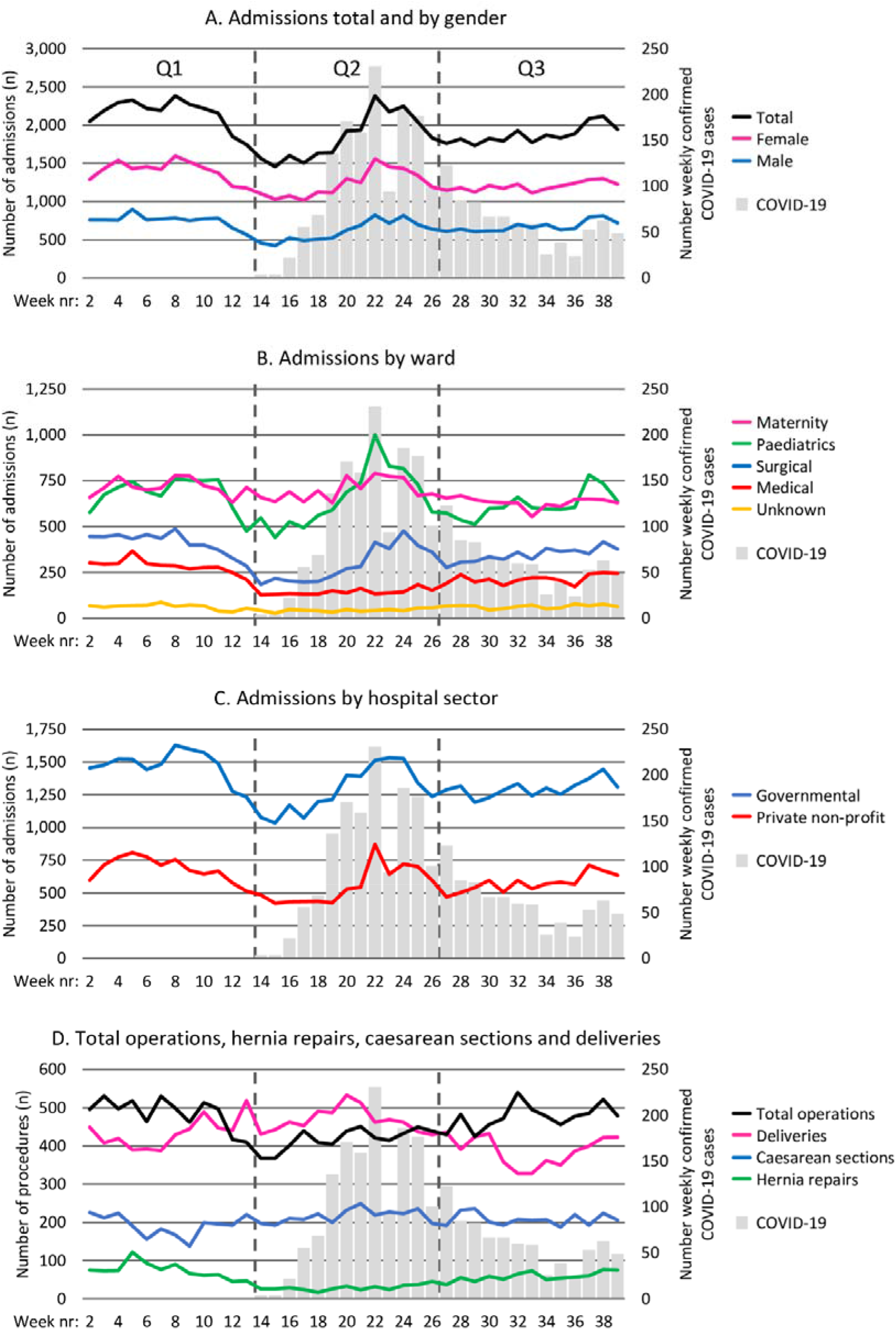
Nationwide mean weekly admissions, disaggregated by sex, overlaid COVID-19 cases, Q1-Q3 2020 Figure 3b: Nationwide mean weekly admissions by service, overlaid COVID-19 cases, Q1-Q3 2020 Figure 3c: Nationwide mean weekly admissions by hospital sector, overlaid COVID-19 cases, Q1-Q3 2020. Figure 3d: Nationwide mean weekly total operations, caesarean sections, hernia repairs and facility-based deliveries, overlaid COVID-19 cases, Q1-Q3 2020.

**Table 2.** Nationwide mean weekly admissions in Sierra Leone, Q1-Q3 2020. *Q1 = 2020 week 2 -13; Q2 = 2020 week 14 – 26; Q3 = 2020 week 27 – 39. *Student t-test*

|  | Q1 | Q2 |  |  | Q3 |  |  | Q2-Q3 |  |
| --- | --- | --- | --- | --- | --- | --- | --- | --- | --- |
|  | Mean (95% CI) | Mean (95% CI) | Q1-Q2 Change % | p-value* | Mean (95% CI) | Q1-Q3 Change % | p-value* | Q2-Q3 Change % | p-value* |
| Total admissions | 2160 (2039 – 2280) | 1842 (1659 – 2026) | -14.7 | **0.005** | 1876 (1804 – 1947) | -13.2 | **<0.001** | 1.8 | 0.715 |
| Admission by sex |  |  |  |  |  |  |  |  |  |
| Male | 753 (703 – 803) | 611 (531 – 691) | -18.8 | **0.004** | 675 (633 – 717) | -10.3 | **0.015** | 10.4 | 0.138 |
| Female | 1407 (1325 – 1489) | 1231 (1125 – 1337) | -12.5 | **0.009** | 1201 (1166 – 1235) | -14.7 | **<0.001** | -2.5 | 0.558 |
| Admissions by ward |  |  |  |  |  |  |  |  |  |
| Paediatrics | 680 (624 – 737) | 657 (560 – 754) | -3.5 | 0.659 | 618 (574 – 662) | -9.2 | 0.067 | -5.8 | 0.435 |
| Maternity | 717 (688 – 745) | 701 (665 – 736) | -2.2 | 0.453 | 632 (615 – 650) | -11.7 | **<0.001** | -9.7 | **<0.001** |
| Surgical | 285 (262 – 308) | 143 (133 – 153) | -49.8 | **<0.001** | 214 (200 – 229) | -24.8 | **<0.001** | 49.8 | **<0.001** |
| Medical | 412 (375 – 449) | 294 (234 – 354) | -28.7 | **0.002** | 346 (323 – 369) | -16.1 | **0.003** | 17.8 | 0.090 |
| Unknown | 63 (54 – 72) | 43 (39 – 48) | -31.0 | **<0.001** | 64 (58 – 70) | 1.8 | 0.817 | 47.6 | **<0.001** |
| Governmental hospitals | 1475 (1400 – 1550) | 1285 (1178 – 1392) | -12.9 | **0.005** | 1300 (1260 – 1339) | -11.9 | **<0.001** | 1.1 | 0.784 |
| Private non-profit | 685 (628 – 742) | 557 (471 – 643) | -18.6 | **0.014** | 576 (535 – 617) | -15.9 | **0.002** | 3.4 | 0.670 |
| Total operations | 486 (461 – 512) | 418 (402 – 435) | -13.9 | **<0.001** | 477 (458 – 496) | -1.9 | 0.526 | 14.0 | **<0.001** |
| Hernia repairs | 74 (60 – 87) | 29 (25 – 33) | -60.7 | **<0.001** | 58 (51 – 65) | -21.0 | **0.030** | 100.8 | **<0.001** |
| Caesarean sections | 192 (175 – 210) | 216 (206 – 227) | 12.7 | **0.014** | 208 (199 – 218) | 8.4 | 0.083 | -3.8 | 0.214 |
| Deliveries | 435 (409 – 460) | 467 (448 – 486) | 7.5 | **0.033** | 388 (364 – 411) | -10.8 | **0.007** | -17.0 | **<0.001** |

From Q1 to Q2 2020 (Figure 3b), mean weekly admissions for surgery decreased from 285 to 143, a 49.8% decrease (p<0.001) and medical admissions decreased from 412 to 294, a 28.7% decrease (p=0.002). Paediatric and maternity admissions did not show a significant change from Q1 to Q2.

In Q3, surgical and medical admissions remained significantly lower than Q1, but with signs of recovery comparing Q3 to Q2 with a 49.8% increase in surgical admissions (p<0.001) and 17.8% increase (p<0.090) in medical admissions. In Q3, maternity services saw a significant decrease compared to Q1, 717 to 632, -11.7% (p<0.001).The reduction in admissions was seen in both the government 12.9% and the private sector 18.6% in Q2, and in Q3, 11.9% and 15.9% respectively, compared to Q1 (Figure 3c).

From Q1 to Q2, total operations decreased from a weekly mean of 486 to 418, a 13.9% decrease (p<0.001), (Figure 3d). Hernia repairs, fell from 74 to 29, a 60.7% decrease (p<0.001). In contrast, caesarean sections and facility-based deliveries showed significant increases, 192 to 216, a 12.7% increase (p=0.014) and 435 to 467 a 7.5% increase (p=0.033). In Q3, there were 477 total operations, no significant difference from Q1 486 (p=0.526), this was through a recovery in hernia repairs, with a 100% increase from Q2 to Q3. Caesarean sections are maintained in Q3 with no significant change. However, facility-based deliveries decreased from 435 in Q1 to 388 in Q3, a 10.8% decrease (p=0.007).

The mean number of referrals per week in Q2 in 2020 was 538 compared to 575 in Q2 in 2019 (p=0.151) (Figure 4). In Q3, the mean number of referrals was significantly lower in 2020 (419) compared to 2019 (530) (p<0.001).

**Figure 4:**
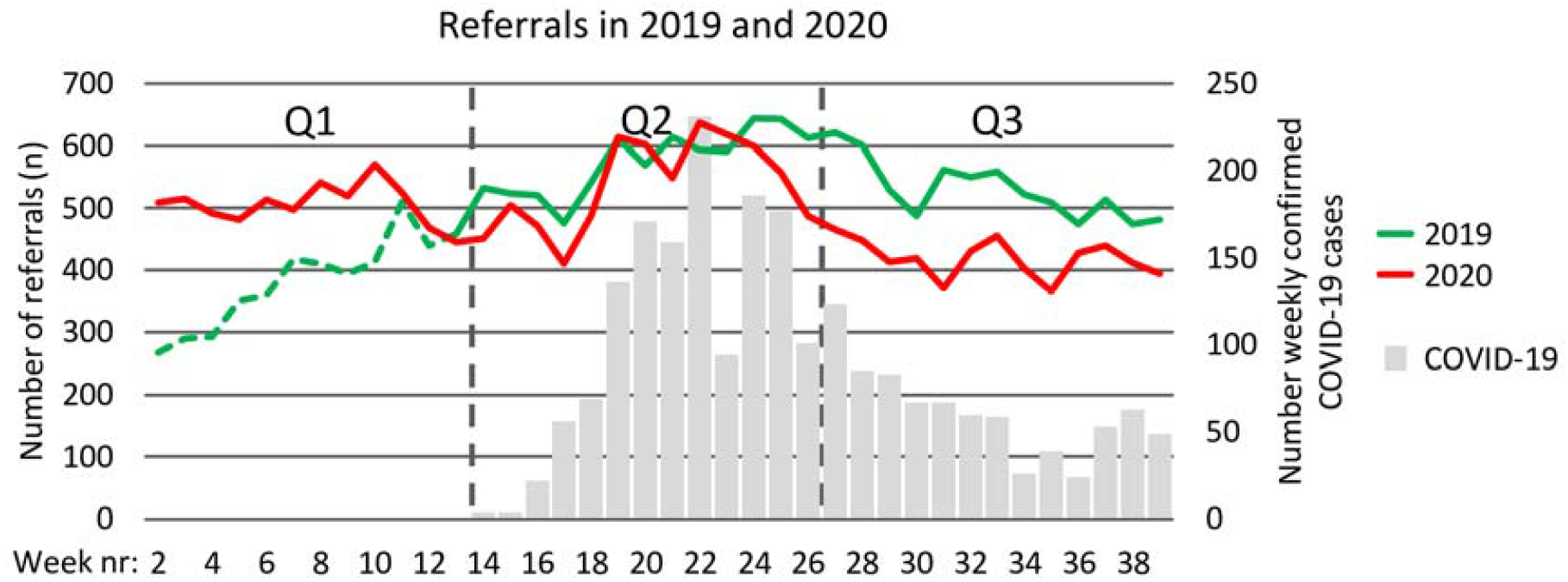
Weekly count of nationwide referrals 2019 and 2020. *The green dotted line represents the start-up phase of the referral coordinator system in 2019

### Factors influencing the provision and utilisation of health services

From the qualitative analysis, we identified both supply and demand-side factors that influenced the provision and utilisation of health services during COVID-19. These factors included *prioritisation of emergency and essential services, introduction of COVID-19 services, and an increase in staff workload* followed by a *return to normalcy*. Demand-side factors included *fear among patients* and *financial barriers*.

#### a) Supply-side changes in health service provision

Respondents reported that the health system response changed over time. Initially, and at the height of the pandemic, there was a planned *prioritisation of services*. This was pursued by allowing only emergency surgeries and essential services such as maternal care, while putting on hold all non-essential services including elective surgeries.

> “At the height of the COVID, everyone was afraid. So only emergency patients were operated on in the surgical department. All elective cases were put on hold…Now we are relaxing those restrictions. (Bo, Secondary Hospital, Doctor)

> “We didn’t reduce [staff] or add but the [staff] roster was changed. (Bo, Secondary Hospital, Maternity Ward Matron)

This adaptive change aimed to prioritise limited resources as well as to limit COVID-19 exposure to non-COVID-19 patients. There was a reduction in bed capacity which aimed to reduce the number of non-COVID-19 patients in the facility and enable social distancing measures.

> “The spacing of the patients’ bed in the ward. Beds were reduced. Yes, because Covid doesn’t favour crowded places. So, beds were being reduced to keep patients apart” (Bo, Secondary Hospital, Doctor).

Our respondents also highlight the prompt *introduction of COVID-19 services*, with the establishment of COVID-19 triage and isolation units for suspected cases. The introduction of these new routines aimed to identify COVID-19 positive patients and reduce nosocomial infection.

> “at the laboratory we have a COVID response team, that does the testing of our patients and the team comes every day for surveillance to check our patients, to take samples, send them to the lab and the lab communicates with the central team and gives us early results of the test” (Western Area Urban, Tertiary Hospital, Surgeon)

However, this caused delays

> “suspected cases based on the symptoms … go to the IDU [Infectious Diseases Unit]. That’s when they do the test… the delay doesn’t really happen at triage. I think, where they move the patient to the IDU, the isolation unit, that [is] where the delay is because we don’t take the samples early and we don’t see the result early enough” (Western Area Urban, Tertiary Hospital, Doctor)

However, some of the respondents also described unplanned service disruption, such as closures of other health facilities

> “[patient numbers] increased. The reason why it increased, we had a positive case at [nearby facility] in the maternity section. So, they closed the theatre and all [services], patients come here now…so, instead of [patients] reducing, it increased… it was too hard, it was overwhelming. (Bo, Secondary Hospital, Maternity Ward Matron)

Our respondents further highlight the speedy *return to normalcy*. However, getting back to normalcy increased patient loads, making the triage system unfeasible

> “The workload increased to an extent that we stopped doing tests. At the beginning, we say every patient that comes, whether it emergency or not, we have to do COVID test. But later, we stopped doing COVID tests for emergency cases because the workload is too much. Waiting for 2 or 3 days, the patient will die” (Western Area Urban, Tertiary Hospital, Surgeon)

#### b) Health seeking behaviour

Beyond the supply-side factors, the reduction in utilisation of non-COVID-19 services in the initial phase of the outbreak was also because of *fear among patients* who were reluctant to come to health facilities for non-emergency care.

> “The influx of patients at the initial stage of the outbreak, dropped. This was because people were afraid to come to the health facility… So that was what led to the low turnout of patients” (Western Area Urban, Primary Health Unit, Deputy In-Charge)

This patient reluctance was increased in relation to facilities that had a COVID-19 treatment unit.

> “Effect of COVID is very severe in this hospital because of the reduction in the number of patients, because patients generally consider that [this hospital] is the centre of COVID” (Western Area Urban, Tertiary Hospital, Surgeon)

Patient’s also experienced *financial barriers*. Respondents reported the cost of transport to health facilities, which increased twofold, and combined with the overall increase in other household expenses and income shocks, added to their financial burden, resulting in reduced healthcare utilisation.

> “So now because of the COVID, they have made [transportation fee] to Le 3000 [instead of the usual Le 1500] … the cost of living is getting higher, more especially the things they are selling, rice and other commodities…I was working but due to the corona, I lost my job, all of us. Since corona, things have not been easy…business too is not easy…the hardship…my brothers who were working, are not working anymore” (Western Area Urban, Patient)
>
> “Well, there was no transportation, especially in district lockdown, transportation was costly, and the people with money travels only… The transport fare from here [home] to [tertiary hospital], they were asking for Le 150,000. Can you imagine? … and [no public transport] so the private vehicles that were going… So, transportation was a serious problem.” (Bo, Secondary Hospital, Patient).

## Discussion

Using nationwide data, we report a significant decrease in hospital admissions after the first COVID-19 case was reported in Sierra Leone, with the decrease continuing into Q3 of 2020. As observed during the Ebola epidemic(16), reductions in healthcare utilisation were not equally distributed across patient groups, services and sectors. Adult medical and surgical services saw the largest decreases, maternal admissions were maintained in Q2 but decreased in Q3, and paediatric admissions showed no significant change throughout the study period. Our findings of a significant reduction in referrals to hospital between Q2 and Q3 in 2020 compared with 2019 suggest that the effects that we show on hospital admissions are unlikely to be seasonal.

The decreases in hospital admissions are lower than reported in Sierra Leone during Ebola. During Ebola, weekly admissions decreased by 51%(16), compared to decreases of 14.7% (Q1-Q2) and 13.2% (Q1-Q3) in this study. The decreases are also lower than the median decrease in admissions of 28.4% (17.4-40.4) from 43 studies in a recent worldwide systematic review(4). We observed similar patterns of change in healthcare utilisation to those found during Ebola. From Q1-Q2 we observed larger decreases in male admissions -18.8% compared to -12.5% in female admissions vs 55% to 50% during Ebola(16). We observed larger decreases in admissions at private non-profit hospitals -18.6% compared to -12.9% at government hospitals vs -60% and -45% during Ebola(16). Total operations decreased from Q1 to Q2, with large decreases in hernia operations and the maintenance of caesarean sections similar to Ebola(16). In contrast to findings from Ebola, we found no significant change in paediatric admissions(2), which may be due to the relatively low child mortality of COVID-19 compared to Ebola(21). Overall, the decreases in utilisation are lower than current reports on COVID-19 indirect effects in other countries(4) and modelled projections(8).

We seek to explain the causal mechanisms of the reductions in utilisation. We posit that the decrease in healthcare utilisation is proportional to the perceived size of the threat. This is supported by the finding that during the 2013-2016 Ebola outbreak, the largest decreases in service utilisation were seen in the districts with the highest Ebola incidence(15). Unlike Ebola, COVID-19 control measures were introduced promptly, the number of cases and reported case fatality remained low(1) and the public perception of the disease was that it was not a serious life-threatening infection. These measures and perceptions were naturally informed by Sierra Leone’s previous experience with Ebola, and many healthcare workers commented on the high mortality rate of Ebola compared to COVID-19 during the interviews. Therefore, COVID-19 was not seen as an existential threat of the same magnitude as Ebola. Aside from the size and perception of the epidemic, decreases in health service utilisation might be a result of: a) adaptive, planned health service reconfigurations; b) unplanned service disruption; or c) changes in health-seeking behaviour.

Planned adaptive service reconfiguration is a deliberate calculated process that anticipates the threat to the health system and adapts to mitigate the threat. A number of planned adaptive health system reconfigurations were made throughout the first six months of the response, leveraging knowledge and experience from the Ebola epidemic. Preparedness plans, policy, ambulance services, coordination and command and control structures were rapidly established and modelled on some of the experiences from previous Ebola response model(24). A National COVID-19 Emergency Response Centre was set up and the response was cascaded to the districts via District COVID-19 Emergency Response Centres. Social mobilisation and community based action groups, which were vital in responding to the Ebola epidemic were reactivated. Cognizant of the previous indirect effects of Ebola on broader health outcomes, the overarching strategic aim of the response was “*saving lives and saving livelihoods*”. Accordingly, the response incorporated the maintenance of non-COVID-19 essential health services as a core objective. To decrease fears of nosocomial transmission, COVID-19 care facilities were clearly delineated from non-COVID care. Case definition-based screening at the front gate of hospitals with linkage to hospital isolation units, provided further delineation between COVID-19 and non-COVID-19 care. Specific ambulances were dedicated solely for COVID-19 and the rest of the fleet maintained for essential health services(25). Rapid expansion of COVID-19 treatment beds was achieved by converting existing hospital spaces into safe treatment and isolation centres, using pre-existing infrastructure and staff and modelled on a previous Ebola response model(26). The response also differed from the Ebola response, in that COVID-19 treatment centres were all located at government hospitals, as opposed to temporary NGO-run separate facilities. We believe this distinction prevented the migration of healthcare workers, retaining them at government facilities where they could fulfil a dual role of delivering COVID-19 and other essential health services. However, this approach of converting normal hospital wards may have also decreased bed capacity for non-COVID-19 care. Healthcare worker incentives for COVID-19 response were also calibrated in an attempt to prevent the pull of healthcare workers away from their normal roles in providing essential services. Additionally, to motivate and engender trust, the health workforce was pro-actively engaged and trained in case management and infection prevention and control(27), and a health and life insurance scheme was introduced for all government healthcare workers. This planned adaptive health system response may have resulted in minimising the indirect effects on health care supply compared to the Ebola epidemic.

The first case management guidelines were published in April 2020 in Sierra Leone. As in many countries, the original policy was to postpone all elective surgery(14), to create extra bed capacity and reduce opportunities for nosocomial transmission. This policy may be partially responsible for the decreased number of elective hernia operations in Q2. Hospitals initially adopted policies of requiring patients to have a negative COVID-19 test before all surgery, with the exception of caesarean sections. Delays in collecting and receiving test results may have acted as a significant disincentive for patients and providers to operate, as was seen during Ebola(28). In anticipation of the first wave of COVID-19, medical superintendents were instructed to create spare bed capacity to deal with the expected incoming surge of COVID-19 patients. This may have led to hospitals imposing stricter admission criteria, reducing the number of both medical and surgical admissions in Q2.

Unplanned services disruption can occur due to health worker infections, industrial actions among health workers, or emergent, unintended consequences of outbreak response interventions or policy. Examples of unplanned service disruption that Sierra Leone has witnessed during the current COVID-19 pandemic include the closure of hospitals, staff infections or quarantine, and healthcare worker strikes(29). In particular, the lack of nuance in applying contact tracing and quarantine procedures for essential healthcare workers at the beginning of the response caused significant unplanned service disruption, leading to closure of hospitals and operating theatres(29). Later, the response developed specific guidance on healthcare worker quarantine and guidance on contact tracing in hospitals, that largely averted further hospital closures. A Doctors strike in July 2020, over delayed payment of COVID-19 incentives, may have affected service delivery. Whilst the strike was focused on COVID-19 care delivery at hospital isolation centres, this widely publicised announcement we believe may have had greater knock-on effects for care-seeking for essential health services. A newly established National Referral Service may have mitigated some of these effects, diverting patients to functioning hospitals, and real time availability of bed capacity across the system. The closure of private hospitals may explain the rise of facility-based deliveries and caesarean sections in Q2 as these services were displaced from the private for-profit sector into governmental hospitals. A similar trend was seen during Ebola, where a 43% decrease in weekly median caesarean deliveries was seen in the private sector, mirrored by an increase of 45% in the government sector(16).

Changes in healthcare utilisation can occur via changes in health-seeking behaviour, through trust in the healthcare system. Trust is affected by patient and community perception of nosocomial infection risk, service availability and quality, as well as cost(7). Patients and communities actively engage in risk benefit decision making when seeking healthcare in Sierra Leone(30). The trend in decreasing admissions begins in week 11 of 2020. This is before the first case was reported in Sierra Leone and closer to the time the first case was recorded in Nigeria (28^th^ February), and before the dissemination of Sierra Leonean guidance or policy. This might suggest that patients and providers began to act independently before official guidance was published. Therefore, a substantial proportion of change in service utilisation was organic and led at the patient and provider level, rather than as a result of a planned national health system reconfiguration.

During the Ebola outbreak, healthcare facilities were correctly identified by patients as “hotspots” for disease transmission(31) and this was a major driver behind decreased healthcare utilisation. In our qualitative study we also find that during COVID-19, patients were reluctant to visit health facilities, particularly large tertiary centres, for this reason. In March 2020, before the virus had arrived in Sierra Leone, a nationwide survey demonstrated high awareness of COVID-19 amongst the public, and the perception of it as a life threatening disease(32). However, as case fatality of COVID-19 remained low at 2.8%, we believe public perception shifted to view it as a less threatening illness. Case fatality rate for COVID-19 must also be weighed against other health threats in Sierra Leone. When compared to an under-5 mortality rate of 12.2% and a maternal mortality rate of 0.7%(33), the risk of contracting COVID-19 when visiting a health facility may be outweighed by the risk of not seeking care(28). This may account for some of the initial drop off in hospital admissions followed by the recovery and resilience of certain sectors of the health system found in our study.

Financial barriers are some of the principal barriers to accessing care in Sierra Leone, especially amongst the non-free healthcare population(34). During the epidemic, people suffered income losses along with a steep rise in transport costs, further increasing the financial barriers to accessing healthcare(35). This could explain why adult medical and surgical admissions, that require more out of pocket expenditure, saw reductions whilst services provided under the Free Health Care Initiative(36) such as caesarean deliveries, were resilient. Furthermore, as the national ambulance service(25) is primarily intended for paediatric and maternal cases, which were unaffected from Q1-Q2, it is possible that the protective effect of free pre-hospital transport system maintained access for these populations.

## Limitations

Our data collectors endeavoured to collect complete admissions data and triangulated with other sources of data in the hospital. However, it is still possible that not all admissions were recorded or that operations or deliveries could have been missed. Our admission data does not include the largest paediatric hospital in the country, as it does not perform surgery, which was severely affected by unplanned service disruption. The absence of admission and surgical activity data from 2019 makes it difficult to assess and adjust for seasonality in our results. We have attempted to account for seasonality and triangulate our data by utilising the National Referral Database, however, it should be noted that these are two distinct data sources. Qualitative data examined health-seeking behaviour but since we interviewed only those people who sought care, we miss experiences of people who did not seek care and who are also more likely to have faced greater barriers. Our study only analyses hospital level data and further research should evaluate primary healthcare utilisation.

## Conclusion

Our study demonstrates a decrease in health service utilisation coinciding with the onset of the COVID-19 pandemic in Sierra Leone. The decreases in health service utilisation are less than seen worldwide, and less than observed during Ebola. The incorporation of maintenance of essential health services as an explicit aim of the outbreak response strategy may have mitigated larger decreases in healthcare utilisation. The resilience of services supported by the Free Health Care Initiative adds further weight to the argument to expand Universal Health Coverage in Sierra Leone. We recommend regular monitoring of health service utilisation in epidemics to guide and evaluate public health response measures.

## Data Availability

Data is avaialble from the corresponding author upon reasonable request, academic approval and acceptance of a data sharing agreement.

## Acknowledgments

We would like to thank all Surgical Assistant Community Health Officers and Research Assistants for their involvement in the project. We are grateful to Professor Foday Sahr, Dr Mathew Vandy and Dr Suliaman Lakoh for their tireless work and their guidance throughout the pandemic. Finally, we express our gratitude to all frontline workers and communities who led the effort to combat COVID-19 in Sierra Leone.

## Competing Interests

No competing interests to declare

### Glossary of abbreviations

COVID-19: Coronavirus disease
IDU: Infectious Disease Unit
LMICs: Low and middle-income countries
sSA: sub-Saharan Africa
Q1, Q2 and Q3: First quarter, second quarter and third quarter
RC: Referral Coordinator

## Notes

### Competing Interest Statement

The authors have declared no competing interest.

### Funding Statement

This research was part funded by the non-profit organisation CapaCare, and part funded by the National Institute of Health Research (NIHR) Global Health Research Unit on Health System Strengthening in Sub-Saharan Africa, Kings College London (GHRU 16/136/54) using UK aid from the UK Government to support global health research. DY is funded by the National Institute for Health Research (NIHR) (GHR:17:63:66) using UK aid from the UK Government to support global health research. The views expressed in this publication are those of the author(s) and not necessarily those of the NIHR or the UK government.

### Author Declarations

Ethical approval was granted by the Sierra Leone Ethical and Scientific Review Committee and the Regional Committee for Medical and Health Research Ethics in central Norway (2020/155388).

